# Polygenic Scores for Dizygotic Twinning: Insights into the Genetic Architecture of Female Fertility

**DOI:** 10.1101/2024.12.02.24318308

**Authors:** Nikki Hubers, Christian M. Page, René Pool, Hamdi Mbarek, Nils Lambalk, Velja Mijatovic, Lannie Ligthart, Jenny van Dongen, Elizabeth C. Corfield, Jeffrey J. Beck, Erik A. Ehli, Nicholas G. Martin, Gonneke Willemsen, Siri Håberg, Jennifer R. Harris, Jouke-Jan Hottenga, Dorret I. Boomsma

## Abstract

Natural dizygotic twinning (DZT) results from hyper-ovulation and is considered an indicator of female fertility. DZT has low polygenicity, with only 0.20% of SNPs estimated to have a nonzero effect. A polygenic score (PGS) for DZT explains 1.6% of variance in DZT liability and we observe an odds ratio of 2.29 between the 1^st^ and the 10^th^ PGS decile. The PGS is higher in mothers of naturally conceived twins compared to mothers who received fertility (MAR) treatments in both the Netherlands and Norway. The largest differences were observed for mothers who received hormonal ovulation induction, indicating maternal fertility issues. A higher PGS was also linked to shorter time to pregnancy. DZT showed significant negative genetic correlations with anovulatory infertility (r_g_ = -0.698) and PCOS (r_g_ = -0.278), and a positive genetic correlation with endometriosis (r_g_ = 0.279). These findings suggest DZT PGS is an important, yet under-recognized fertility marker.

## Main

Giving birth to naturally conceived dizygotic (DZ) twins starts with poly-ovulation, followed by multiple fertilization and successful implantation of at least two embryos (1). DZ twinning (DZT) occurs more often in some families than in others and is partially genetically influenced through the mothers’ side, which is what we would expect for a trait that is dependent on the mother’s physiology (1–4). Two genome-wide association studies (GWAS) report multiple independent loci associated with DZT (5,6). The SNP-based heritability was estimated to be 2.4% on the liability scale in the most recent GWAS meta-analysis of DZT (6). The heritability estimate for DZ twinning is 28%, based on a reanalysis of sister-pair data (4). An analysis of seven large multi-generations pedigrees from West Africa, fin-de-siècle French Jewish populations, Canadians and the French royal family obtained an estimate of 8 to 20%, without distinguishing between mono- and dizygotic twins (4,7).

DZT presents as an ‘atypical’ complex trait in genetic studies. The phenotype definition is critically important as illustrated by the first GWAS for DZT, when a genetic signal was observed only after removing a small subset (N = 241) of mothers of DZ twins who had conceived twins after medically assisted reproduction (MAR) (8). Such a strong effect of phenotype misclassification hints at opposing genetic effects associated with the natural conception of DZ twins versus mothers with subfertility or infertility. Also, in the most recent meta-analysis, the genomic variants which were identified generally are located in the coding regions of the genome, as opposed to non-coding regions that are often found in GWAS (9,10). Predicted causal genes are involved in endocrine processes of female fertility including follicle stimulating hormone subunit B (*FSHB*), gonadotropin releasing hormone 1 (*GNRH1*), and follicle stimulating hormone receptor (*FSHR)* (6,11).

It has been proposed that DZT can be viewed as an indicator of high female fertility and that research into DZT can thus aid in understanding female fertility and infertility in general (12) Paradoxically, several characteristics associated with DZT are associated with infertility (13–15). Infertility is defined as being unable to conceive after 12 consecutive months of trying and has been estimated to affect 1 in 6 couples world-wide (16). Several underlying conditions may lead to female infertility including polycystic ovary syndrome (PCOS) and endometriosis, but for at least 15% of couples the underlying reason for infertility remains unclear (17). MAR treatments for infertility have been available since the late 1960’s and can result in increased dizygotic twinning rates for two reasons: 1) increase in hyper ovulation as a result of hormonal ovulation induction (OI) mimicking the natural DZT mechanism (1,18) or 2) the transfer of multiple fertilized oocytes after in-vitro fertilization (IVF) cycles to the uterus, although the transfer of more than one fertilized oocyte has become less common in recent years (19,20).

Our objectives were to investigate the genetic architecture of DZT and to test the hypothesis that DZT can be regarded as an indicator of female fertility. We first investigated the polygenicity of DZT by employing multiple tools (21–24). Next, we compared the polygenic score (PGS) for DZT between twin and singleton mothers who conceived naturally and mothers who received MAR treatments. We tested our hypothesis in the Netherlands Twin Register (NTR) and in the Norwegian Mother, Father and Child Cohort Study (MoBa). We extended the PGS analyses by assessing whether the DZT PGS was associated with the time to achieve pregnancy in singleton mothers. To further understand the relation between (sub)fertility and DZT we estimated the genetic correlations between DZT and seven fertility phenotypes (21,22).

## Results

### Polygenicity of DZ Twinning

To investigate the polygenicity of DZT we used two genetically informed tools. We employed GENESIS (https://github.com/yandorazhang/GENESIS) for predicting the genetic effect-size distribution based on the summary statistics of the 2024 GWAMA of DZT (6,23). We fitted both the M2 and M3 models that GENESIS provides. Figure S1 shows that the M3 model was best suited implying that DZT is influenced by a mixture of SNPs with a small effect and a larger effect. We calculated an effective sample size of discovery GWAS for DZT by the described formula: (8,265 + (26,252 / 2)) x 682,000 / 716,517) = 20,361. Mothers of DZ twins are (N = 8,265) counted as one case while DZ twins (N = 26,252) are counted as half cases given that the twins carry half of their mothers genes. The total predicted number of susceptible SNPs for DZT is around 2,244 (SD = 1,498) and the predicted maximum SNP heritability based on these 2,244 SNPs was 7.2% (SD = 2.4%). Of the 7.2% SNP heritability, 17.2% (SNP heritability = 1.2%; SD = 0.6%) is predicted to be explained by a cluster of only 17 SNPs with the largest effect size. The results of GENESIS also show that a sample size of approximately 400,000 participants (200,000 cases and 200,000 controls; Figure S2) would be required to identify 80% of the total SNP-based heritability.

Next, we ran SbayesS to evaluate the polygenicity of DZT. SbayesS estimated the proportion of SNPs with nonzero effects as an indication of the polygenicity (24)https://cnsgenomics.com/software/gctb/). We used a LD matrix based on UK Biobank (UKB) provided on the SbayesS webpage (https://cnsgenomics.com/software/gctb/#Download) and the summary statistics of the DZT GWAS by Mbarek et al., 2024 (6). SbayesS estimated that about 0.198% (SE = 4.00x^-5^), or approximately 5,483 (SD = 939) of SNPs will have a nonzero effect with regard to DZT. The two estimates of the susceptible SNPs from GENESIS and SbayesS are not significantly different from each other based on the surrounding confidence intervals, indicating robustness in the estimates. SbayesS estimates the SNP based heritability of DZT based on the 2024 GWAS at 0.8% (SE = 9.19x^-5^).

### Variance explained by the DZT PGS

To investigate the optimal fraction of SNPs to include in the PGS, we calculated the explained variance in NTR on the liability scale in a sample of 1,635 natural mothers of DZ twins and 2,797 controls. We first calculated PGS using LDpred 0.9 softwareV1 (Methods) with several fraction of SNPs (*f*) with non-zero effects (infinitesimal, 0.5, 0.3, 0.2, 0.1, 0.05, 0.01, 0.005, 0.003, and 0.001). Figure 1 shows that explained variance range of 0.71-1.60% on the liability scale (Table S1). The highest variance explained was observed for *f* = 0.001 and thus, this was the fraction selected to calculate the PGS in NTR and MoBa. Figure 1 also includes a PGS solely based on 26 genome-wide significant SNPs which in the 2024 GWAS were included in a PGS to explore global differences in DZT. Clearly this PGS had the lowest explained variance (R^2^=0.71%), pointing to a need for more research in non-EU populations.

**Figure 1:**
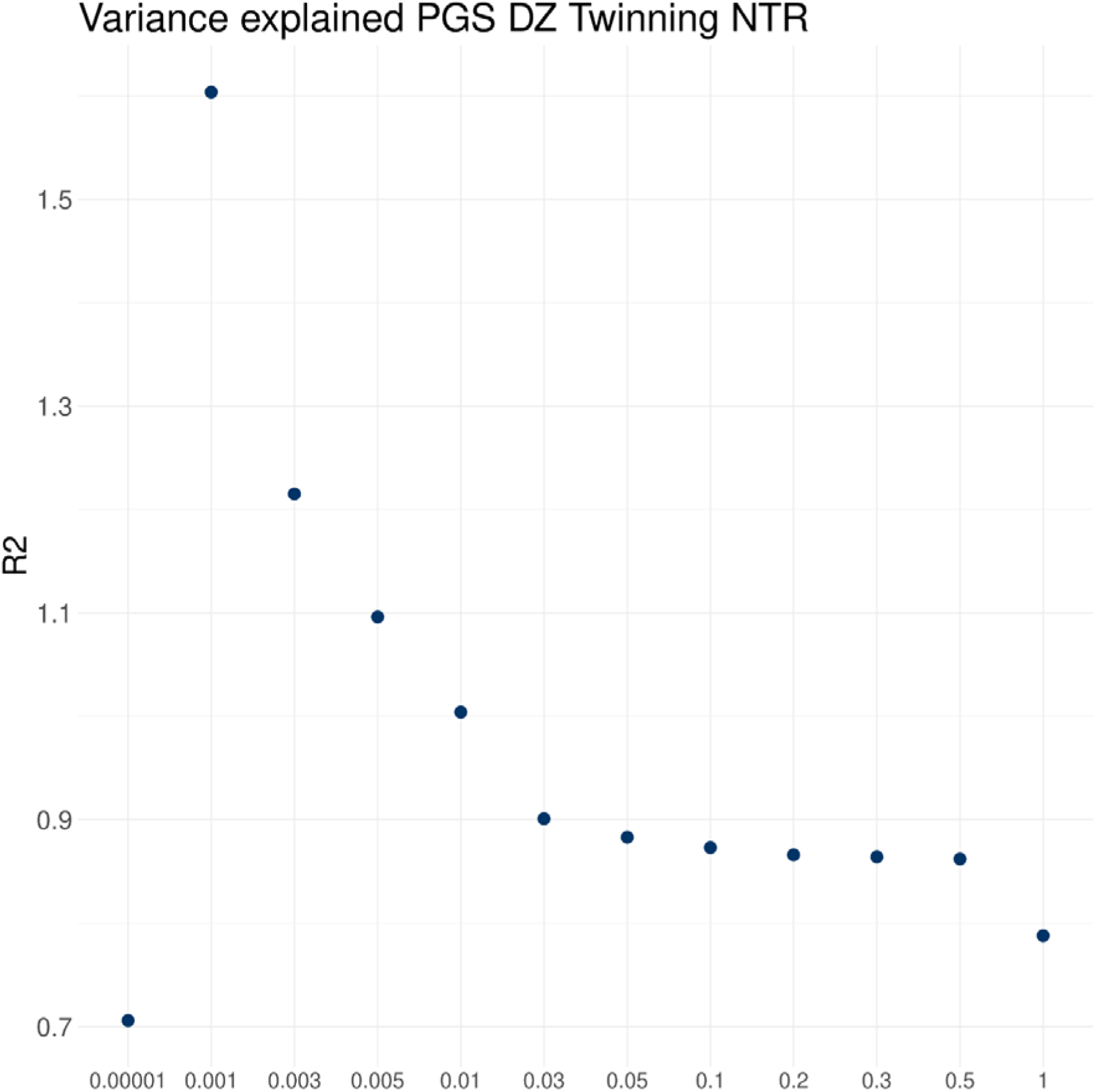
The variance explained on the liability scale by the polygenic score (PGS) for dizygotic (DZT) twinning in 1,635 cases and 2,797 controls for each fraction of top SNPs from the 2024 DZT GWAS included in the PGS.

### PGS analyses

#### Odds ratios

The PGS of all 4,381 mothers of twins and 2,797 controls in NTR were and split into 10 decile groups. We calculated odds ratios for giving birth to a naturally conceived DZ twin and giving birth to a DZ twin after MAR treatments in each decile compared to the 1^st^ and the 5^th^ decile. Figure 2A summarizes the odds ratio compared to the 5^th^ decile, indicating a 1.67 increased risk of giving birth to a DZ twin in the 10^th^ decile compared to the 5^th^ decile (P = 2.91x^-5^). Between the 1^st^ and 10^th^ decile, the increase in risk was 2.29 (P = 1.60x^-10^). Figure 2B indicates an 1.58 increased risk for needing MAR treatments to conceive a DZ twin in the 1^st^ decile compared to the 5^th^ decile (P = 0.024) and a 1.54 increased risk in the 2^nd^ decile compared to the 5^th^ decile (P = 0.034). The odds of needing MAR treatments were not increased between the 1^st^ and 10^th^ decide.

**Figure 2:**
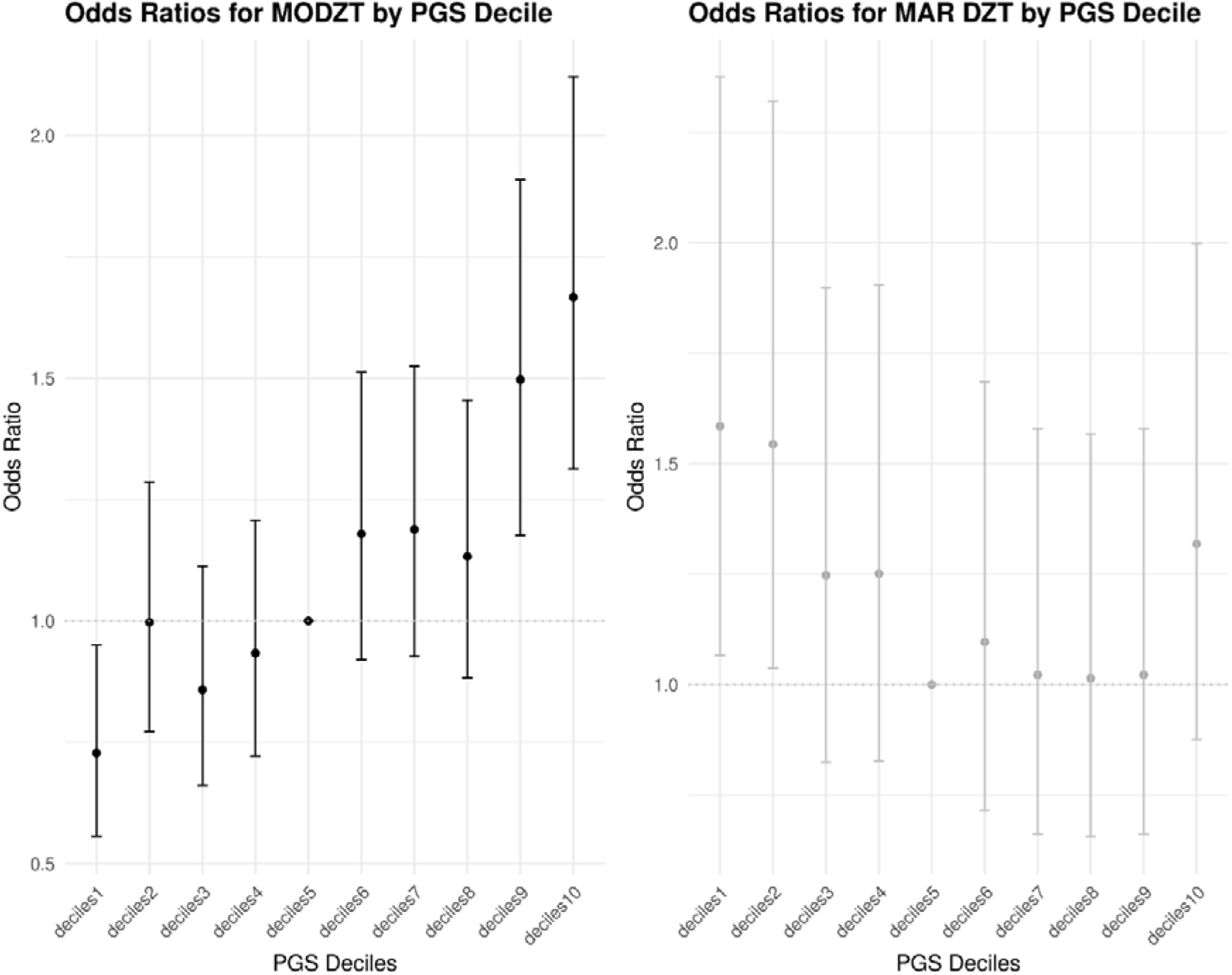
Odds ratios for dizygotic twinning (DZT) in the 7,178 Netherlands Twin Register (NTR) participants by PGS decile. Odds ratios and 95% confidence intervals were estimated with logistic regression for both **A)** being of mother of a naturally conceived DZ twin and **B)** being a mother of a DZ twin who conceived after medically assisted reproduction (MAR). The reported odds ratios are relative to the fifth decile. The points represent odds ratios and the bars represent the lower and upper 95% CI of the odds ratios.

#### Controls and mothers of naturally conceived DZ twins

We compared the DZT PGS between mothers of natural DZ twins and controls from the NTR using the PGS based on the *f* = 0.001 of the SNPs with a non-zero effect. Table 1 and S2 and figure 3A show that the mean PGS was significantly higher in the mothers of naturally conceived DZ twins compared to the controls (P = 6.94x10^-13^). In MoBa, the mothers of natural DZ twins also had a significantly higher mean PGS for DZT compared to controls (the mothers of naturally conceived singletons; P = 1.20x10^-4^; Table 2 & S3; Figure 3B).

**Figure 3:**
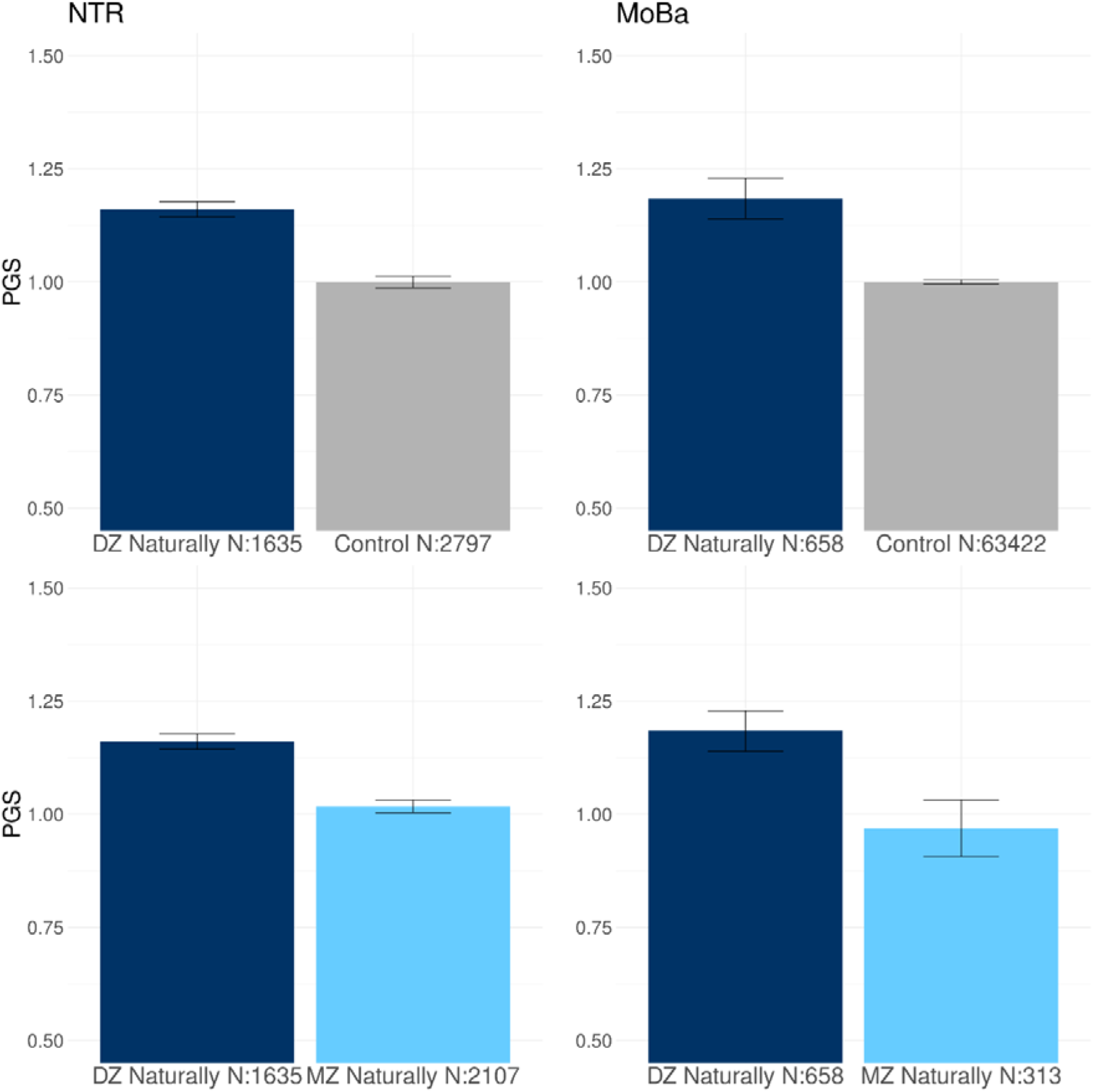
The mean PGS for dizygotic (DZ) twinning in mothers of naturally conceived DZ, controls and monozygotic (MZ) twins from the **A and C)** Netherlands twin register (NTR) and **B and D)** the Norwegian Mother, Children, and Father (MoBa) cohort study. MAR = Medically assisted reproduction.

**Table 1:**
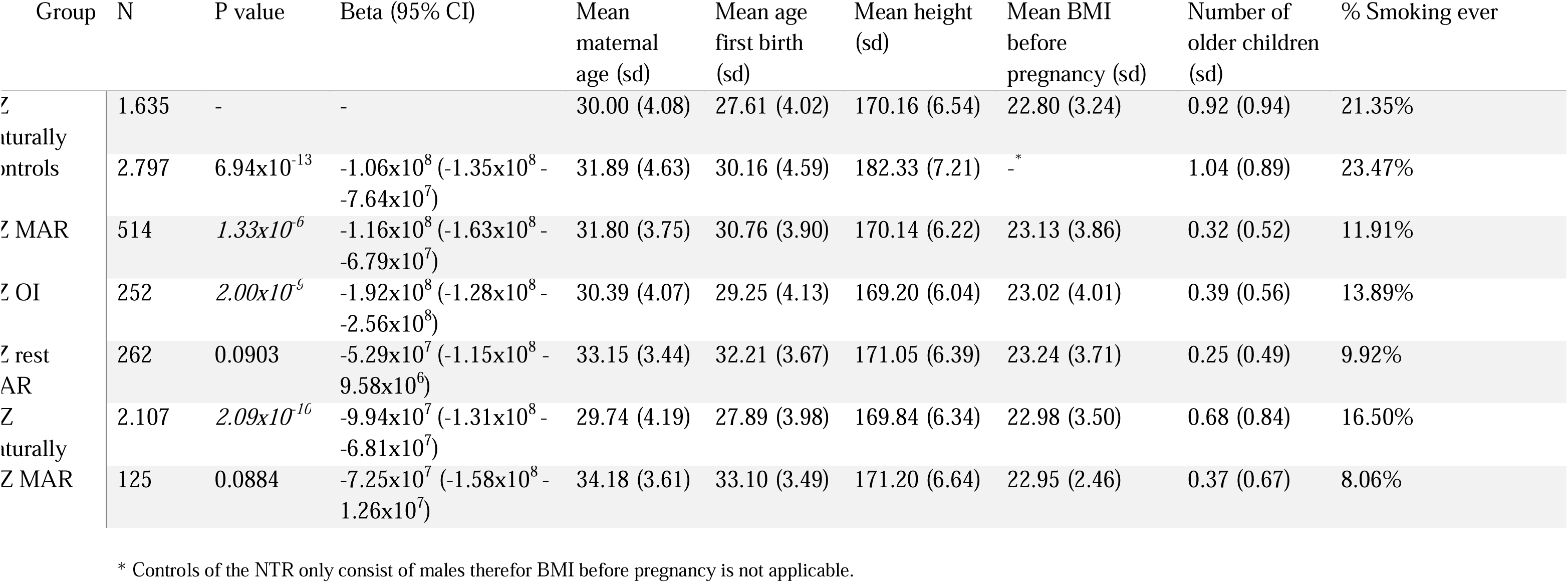
The comparison of the polygenic scores (PGS) for dizygotic (DZ) twinning in mothers of DZ with controls and other mothers of twins from the Netherlands Twin Register (NTR). The controls consist of fathers of twins. All the analyses are shown with the mothers of naturally conceived DZ twins as the reference group. MAR = Medically assisted reproduction; OI = Ovulation induction.

**Table 2:**
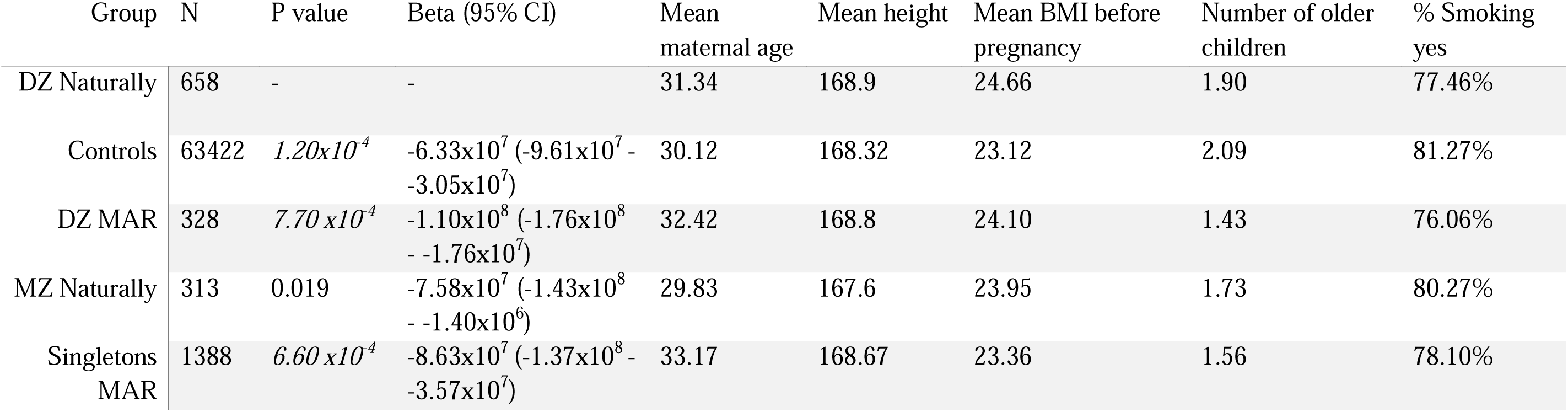
The comparison of the polygenic score for dizygotic (DZ) twinning in mothers of DZ twins with mothers of MZ twins and singletons from the Norwegian Mother, Children, and Father (MoBa). The controls include the mothers of naturally conceived singletons. All the results are shown with the mothers of naturally conceived DZ twins as the reference group. MAR = Medically assisted reproduction.

| Group | N | P value | Beta (95% CI) | Mean maternal age | Mean height | Mean BMI before pregnancy | Number of older children | % Smoking yes |
| --- | --- | --- | --- | --- | --- | --- | --- | --- |
| DZ Naturally | 658 | - | - | 31.34 | 168.9 | 24.66 | 1.90 | 77.46% |
| Controls | 63422 | $1.20 \times 10^{-4}$ | $-6.33 \times 10^7$ ($-9.61 \times 10^7$ - $-3.05 \times 10^7$) | 30.12 | 168.32 | 23.12 | 2.09 | 81.27% |
| DZ MAR | 328 | $7.70 \times 10^{-4}$ | $-1.10 \times 10^8$ ($-1.76 \times 10^8$ - $-1.76 \times 10^7$) | 32.42 | 168.8 | 24.10 | 1.43 | 76.06% |
| MZ Naturally | 313 | 0.019 | $-7.58 \times 10^7$ ($-1.43 \times 10^8$ - $-1.40 \times 10^6$) | 29.83 | 167.6 | 23.95 | 1.73 | 80.27% |
| Singletons MAR | 1388 | $6.60 \times 10^{-4}$ | $-8.63 \times 10^7$ ($-1.37 \times 10^8$ - $-3.57 \times 10^7$) | 33.17 | 168.67 | 23.36 | 1.56 | 78.10% |

#### Mothers of naturally conceived DZ and MZ twins

In NTR we observed a significantly higher mean PGS in the mothers of natural DZ twins than in the mothers of natural MZ twins, for whom their twin pregnancy is not the result of a double ovulation (P = 2.09x10^-10^; Table 1 & S2; Figure 3C). In MoBa the mean PGS is also higher in the mothers of natural DZ twins compared to the mothers of naturally conceived MZ twins based on a nominal p-value of 0.05, but this was attenuated after correcting for multiple testing (P = 0.019; Table 2 & S3; Figure 3D). When comparing controls to mothers of natural MZ twins, we observed no significant difference in the PGS for DZT in the NTR (P = 0.350; Table S4) or MoBa (P = 0.84 ;Table S5; Figure S3A). In NTR, mothers of MAR MZ twins (P = 0.185; Table S4; Figure S3B) did not differ from the controls.

#### Mothers of naturally conceived DZ twins and of DZ MAR twins

Mothers of natural DZ twins have a higher PGS on average compared to mothers of MAR DZ twins (P = 1.33x10^-6^; Table 1; Figure 4A). The significant difference between mothers of natural DZ twins and the MAR DZ twin mothers was also observed in MoBa (P = 7.70 x10^-4^; Table 2; Figure 4B). We then separated the DZ MAR mothers in NTR into a group of mothers who received OI treatment and the IVF/ICSI MAR mothers. Here we observed that the mothers who received OI treatment, where couple infertility likely has a female component, had a lower PGS for DZT compared to the mothers of naturally conceived DZ twins (P = 2.00x10^-9^; Table 1; Figure 5). The PGS in the other groups of DZ MAR mothers, where couple infertility can be due to male, female or combined infertility, did not differ from those in the mothers of naturally conceived DZ twins (Table 1), indicating that the difference in PGS between the mothers of naturally conceived DZ twins and the MAR DZ mothers, is driven by maternal infertility issues.

**Figure 4:**
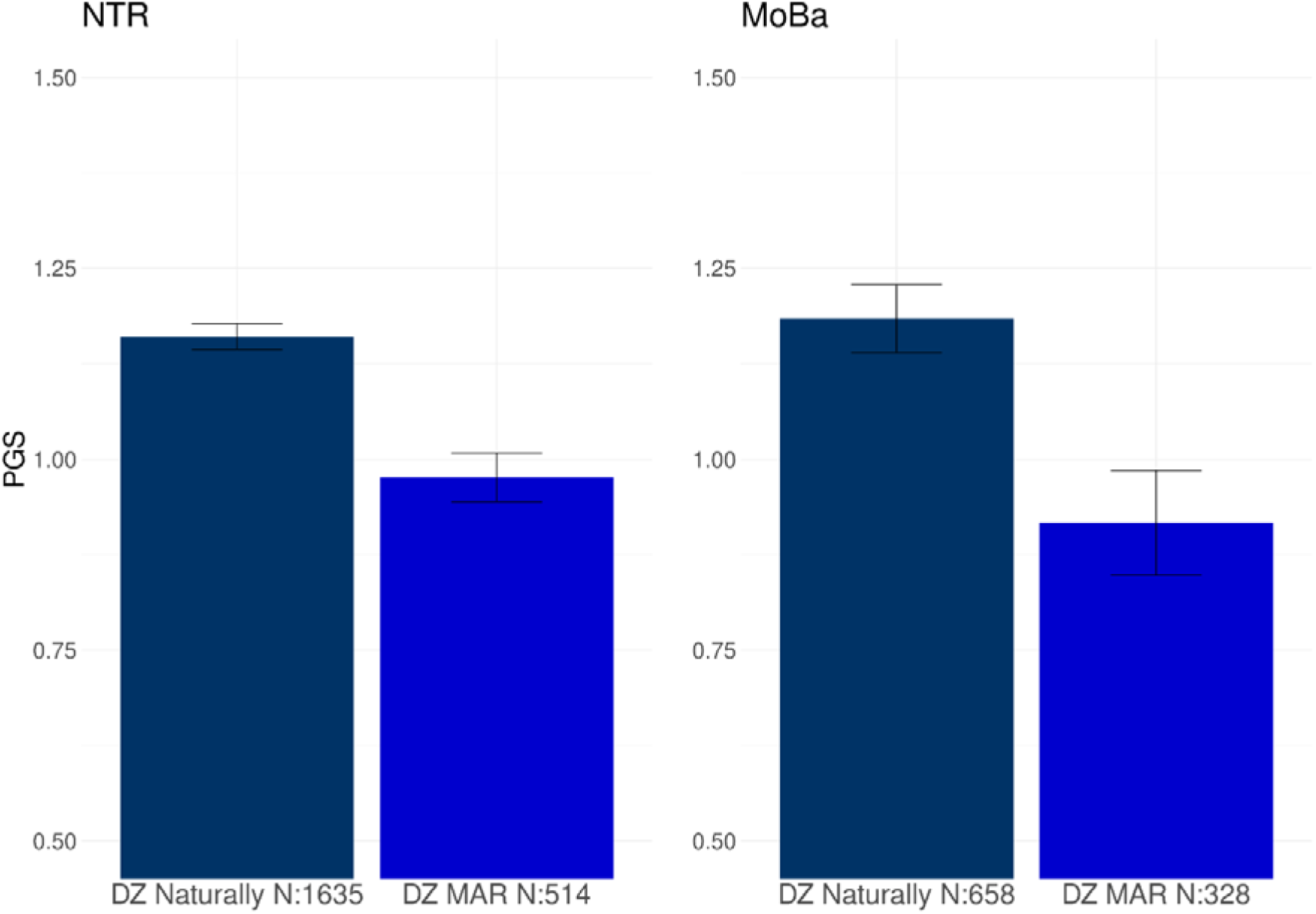
The mean PGS for dizygotic (DZ) twinning in mothers of naturally conceived DZ twins and mothers from DZ twins that received medically assisted reproduction (MAR) treatments from **A)** the Netherlands Twin Register (NTR) and **B)** the Norwegian Mother, Children, and Father Cohort Study (MoBa).

**Figure 5:**
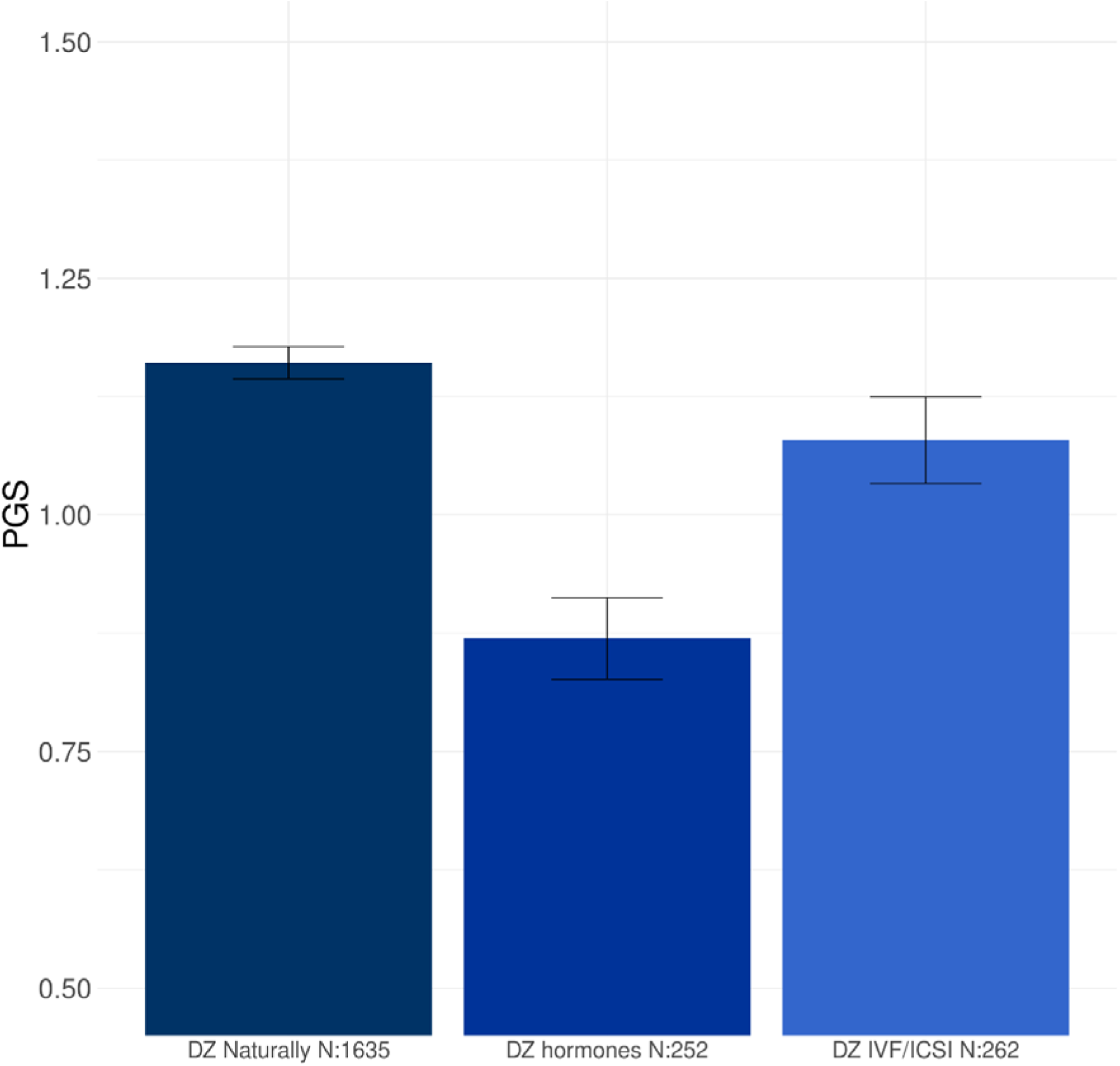
The mean PGS of the DZ MAR group of the NTR is further split between ovulation induction (OI) and remaining MAR treatments.

#### Singleton mothers

No difference was observed between mothers of the natural singletons and MAR singletons in MoBa (P = 0.32; Table S5; Figure S4). The inclusion of covariates, i.e., maternal age, age at first birth, height, BMI before pregnancy, number of older children, and smoking, did not change these results (Tables S2 & S3).

#### Time to pregnancy

In the mothers of naturally conceived twins (NTR) and singletons (MoNa) we performed regression analyses between the DZT PGS and time to pregnancy measured in months. We found no effect in the mothers of naturally conceived twins (Table S6), however, we observed that the DZT PGS was associated with a shorten TTP (P = 0.0036; Table S7) when looking at TTP as a continuous variable in the much larger group of singleton mothers. In a categorial, per month, analysis in MoBa we observed a significant decrease of the PGSs for the mothers for whom it took 12 or more months to conceive compared to mothers who had an unplanned pregnancy (P = 0.0138; Table S7).

### Genetic correlations

We calculated genetic correlations between DZT and seven fertility related phenotypes including PCOS, anovulatory infertility, endometriosis, number of children ever born (NEB), childlessness (CL) and male infertility (Table 3). DZT correlated negatively with anovulatory infertility (r_g_ = -0.698; P = 4.93x10^-5^) and PCOS (r_g_= -0.278; P = 0.0353) and positively with endometriosis (r_g_= 0.279; P = 0.0033).

**Table 3:** Genetic correlations between dizygotic twinning (DZT) and seven fertility related phenotypes. The reference indicates the original publication of the GWAS. r_g_ = Genetic correlation

| Trait | N | SNP based heritability | P heritability | $r_g$ | SE $r_g$ | P $r_g$ | Reference |
| --- | --- | --- | --- | --- | --- | --- | --- |
| DZT | 716.517 (8.265 MODZT, 26.252 DZT) | 0.50% | 0.001 | - | - | - | Mbarek et al., 2024 |
| Anovulatory infertility | 323.473 (7.359 cases) | 0.80% | 0.002 | -0.698 | 0.162 | $5.099 \times 10^{-5}$ | Venkatesh et al., 2024 |
| Polycystic ovarian syndrome | 113.248 (10.074 cases) | 12.40% | 0.020 | -0.278 | 0.132 | 0.035 | Day et al., 2018 |
| Miscarriages | 428.523 (69.054 cases) | 0.43% | 0.001 | -0.165 | 0.151 | 0.272 | Laisk et al., 2020 |
| Endometriosis | 474.160 (60.674 cases) | 1.63% | 0.002 | 0.279 | 0.095 | 0.003 | Rahmioglu et al., 2023 |
| Number of children ever born | 785.604 | 2.11% | 0.001 | 0.093 | 0.064 | 0.147 | Mathieson et al., 2023 |
| Childlessness | 450.082 | 3.62% | 0.002 | 0.014 | 0.073 | 0.853 | Mathieson et al., 2023 |
| Male infertility | 3.225 (1.274 cases) | 11.5% | 0.327 | 0.048 | 0.121 | 0.694 | Cerván-Martín et al., 2022 |

## Discussion

Our results indicate DZT to be a valuable phenotype for studying female fertility and infertility as the DZT PGS distinguishes between twin mothers of naturally conceived pregnancies, and twin mothers who received MAR treatments and can identify those at an increased risk of requiring MAR treatments. The PGS is also associated with a shorter time to pregnancy and is lowest in mothers who received hormonal induction of ovulation, suggesting that the genetic variants associating with DZT primarily affect ovulation processes. This suggestion is supported by the strong genetic correlation between DZT and anovulatory infertility.

We observe that, in contrast to many other human traits, the best performing DZT PGS does not include all possible SNPs (25). Here we include the *f* = 0.001 of the SNPs with a non-zero effect, indicating that DZT is not as poly- or omni genic as other complex traits (26). The GENESIS analysis further confirms that DZT is more likely associated with a smaller number of variants with larger effect sizes as GENESIS predicts 2,244 susceptible SNPs for DZT, while it has predicted over 30,000 susceptible SNPS for highly polygenic traits such as major depressive disorder (23). SbayesS predicts that approximately 0.2% of all SNPs will have a nonzero effect on DZT while for highly polygenic traits such as educational attainment more than 5% of common SNPs are predicted to have a nonzero effects (24). The relatively low polygenicity for DZT found here is in line with findings for other fertility traits in a recent large infertility GWAS (27,28). These results imply that the few variants associated with DZT are likely to have a large effect on overall female fertility.

The PGS based on the first, 2016 DZT GWAS, which obtained evidence for *FSHB* and *SMAD3*, was associated with having children, greater lifetime parity, and earlier age at first child in the Icelandic population (8). Here, we add that the PGS for DZT, based on the 2024 DZT GWAS, is higher in mothers who naturally conceived DZ twins compared to DZ twin mothers who received MAR treatments. In the mothers of singletons the PGS is associated with a reduced time to pregnancy. These findings highlight the connection between DZT and female fertility, emphasizing their potential significance for women contemplating delaying childbirth. While a lower PGS could in future serve as an important indicator for those considering egg freezing at a younger age, it’s important to note that this approach may not yet fully feasible for implementation for this purpose.

The genetic signal detected for DZT is most likely driven by genetic influences on ovulation rate, as the largest difference in PGS was observed between the mothers of naturally conceived DZ twins and the mothers who received OI treatments, which generally indicate maternal fertility issues due to anovulation problems (18,29). Anovulatory infertility is in direct contrast to the hyper-ovulation seen in mothers of naturally conceived DZ twins which is also confirmed with the strong negative genetic correlation between DZT and anovulatory infertility estimated at r_g_ = -0.698 (30,31). These findings point to ovulation rate as a genetic spectrum with anovulatory infertility on one end and hyper-ovulation as indicated by natural DZT on the other end.

Pregnancies after MAR, especially twin pregnancies, carry higher mortality and morbidity risks for mother and child and more miscarriages are seen with MAR treatments (32–34). Also, MAR treatments have on average a low success rate of approximately 20% varying between 5 and 40% depending on e.g. maternal age (19,35). Our results support the idea that genetic investigation with respect to DZT could aid in improving the outcomes of MAR treatments (36). For example, combining the DZT PGS with clinical features may serve in better predictions of who will need MAR treatments and which specific treatments has the best chance of success (37).

Female infertility is an overarching term that covers heterogenous problems. DZT was negatively correlated with anovulatory infertility and PCOS (r_g_ = -0.698 and r_g_ = -0.278 respectively), but positively with endometriosis (r_g_ = 0.279). The positive correlation with endometriosis seems puzzling, but is in line with a recent study that compared pregnancy outcomes among women with endometriosis and uterine cysts and controls which observed that mothers with endometrioses have an increased risk for twinning, regardless of MAR status (38).

It has been proposed that female fertility traits are mediated by a common genetic factor, though the direction of effects of these factors seem to vary per fertility trait (39). A potential common genetic factor is the top locus of the DZT GWAMA, *FSHB* as this locus has also been associated with endometriosis, PCOS, ovarian cyst and menorrhagia and the direction of effect differs between traits (6,39–43).

Some limitations should be considered with our study. The underlying reasons for MAR treatments in NTR and MoBa could be caused by maternal or paternal fertility problems, or a combination of both. Treatment type can serve as an indication of the origin of the fertility problems as OI most likely indicates maternal problems, but we cannot exclude the possibility that reduced male fertility also plays a role. The lack of further differentiations may also explain why no difference is observed in singleton mothers of the MoBa cohort. Still, in singleton mothers, an increased DZT PGS was associated with a shorter TTP, further supporting the relation between DZT and heightened female fertility.

Our sample is based on individuals of European ancestries. DZ twinning is a trait with huge global differences in prevalence (44,45). To test if the effect is also present in other ancestries, we calculated the PGS in three groups of twin mothers in non-European ethnic groups (mostly Caribbean ancestry; N=118) from the NTR. Similar to the main findings based on the sample from European ancestry, Figure S5 suggest that the PGS is lower in the DZ twin mothers who received MAR treatments compared to the mothers of naturally conceived DZ twins.

However, follow-up based on larger sample sizes is clearly required. GWAS and PGS analyses in diverse samples with high DZ twinning rates, e.g. as observed in Nigeria, can enable tests of genetic explanations for the large global differences.

In summary, we find evidence for a genetic spectrum ranging from anovulation/infertility to becoming the mother of spontaneous DZT. A persons liability on this scale informs on DZT risk and other female fertility traits such as time to pregnancy, PCOS and endometriosis and implies that DZT is a valuable model for understanding female infertility.

## Methods

### Participants and phenotypes

#### NTR

The Netherlands twin register (NTR) is a national twin-family register established in the late 1980s (46,47). A total of 4,381 genotyped mothers were included and categorized into four groups: mothers of natural DZ twins (N = 1,635); mothers of natural MZ twins (N = 2,107); MAR MZ mothers (N =125) and MAR DZ mothers (N = 514). MAR was defined as the use of IVF, intracytoplasmic sperm injection (ICSI) and OI. The DZ MAR mothers were further stratified into mothers of DZ twins who received hormonal induction ovulation without IVF (OI; N = 252) and the other MAR DZ mothers (N = 262) who underwent IVF or ICSI. Mothers were excluded from the study if they were not the biological mother (N = 92), if they were a mother of both MZ and DZ twins (N = 65), or if they were mothers of MZ twins and received OI treatments (N = 27). We included 2,797 fathers of twins as controls. Given that the fathers were screened on not being a multiple themselves and that there is no evidence for assortative mating for twinning, no confounding effects were expected by using fathers of twins as controls (48). Mothers without information on MAR were classified as non-MAR when the twins were born before 1970, as IVF was introduced in the Netherlands in the late 1970’s and clomid (a hormonal substitute used to induce ovulation) in the late 1960’s, (49), Surveys provided information on maternal age at birth of twins, age first birth, height, BMI before pregnancy, number of older children, and smoking before the twin pregnancy (15). We determined time to pregnancy (TTP) for 2,298 mothers of naturally conceived twins. TTP was determined in four categories; 0-2 months, 3-5 months, 6-12 months and more then 12 months. The study protocol was reviewed by the medical ethics review committee of the University Medical Centre of Amsterdam (IRB) (Numbers 2003/161 and 548 6565).

#### MoBa

The Norwegian Mother, Father and Child Cohort Study (MoBa) is a population-based pregnancy cohort study conducted by the Norwegian Institute of Public Health (50). Participants were recruited from maternity wards all over Norway from 1999-2008. The women consented to participation in 41% of the pregnancies. Blood samples were obtained from both parents during pregnancy and from mothers and children (umbilical cord) at birth (51). The cohort includes approximately 114,500 children, 95,200 mothers and 75,200 fathers.

The current study is based on version 12 of the quality-assured data files released for research in January 2019. The establishment of MoBa and initial data collection was based on a license from the Norwegian Data Protection Agency and approval from The Regional Committees for Medical and Health Research Ethics. The MoBa cohort is currently regulated by the Norwegian Health Registry Act. This study was approved by the Regional Committees for Medical and Health Research Ethics of South East Norway (#2017/1362). All participants provided written informed consent. The establishment of MoBa and initial data collection were based on a license from the Norwegian Data Protection Agency and an approval from the Regional Committees for Medical and Health Research Ethics. The MoBa cohort is currently regulated by the Norwegian Health Registry Act.

Data on the participating mothers were linked with data from the Medical Birth Registry of Norway (MBRN; a national health registry containing information about all births in Norway), to assess their reproductive history. We included 1,299 mothers of DZ twins and 64,810 mothers of singletons. Mothers were excluded from the singleton group if they had an earlier or later twin pregnancy (N= 2,110). We categorized mothers of twins into three groups: mothers of natural DZ twins (N=658), mothers of MAR DZ twins (N=328), and mothers of natural MZ twins (N=313). Mothers of singletons were categorized as mothers of natural singletons (N=63,422) or mothers of MAR singletons (N=1,388). The mothers of naturally conceived singletons were the control group in the MoBa analyses. Mode of conception was reported by record linkage to the MBRN, which contains information on IVF/ICSI use as reported from the fertility clinics, or by self-report in MoBa. Other information on maternal age, reproductive history and maternal body composition and TTP was obtained from MoBa questionnaires or from the MBRN (15,52). We determined TTP for 42,285 mothers of naturally conceived singletons and TTP ranged between 0 (unplanned pregnancy) till 12 or more months.

### PGS calculation

#### NTR

The complete genotyping and imputation information from the NTR can be found in Data S1. The PGSs in the NTR sample were based on the discovery GWAS meta-analysis for DZT of Mbarek et al. (6). A Leave-one-out (LOO) meta-analysis was performed, removing all NTR results from the meta-analysis and rerunning the meta-analysis as detailed in Mbarek et al. (2024). We retained variants for which the effect allele frequency (EAF) was 0.01 ≤ EAF ≤ 0.99. Variant EAF and effect sizes were aligned with the NTR reference for the 1000 Genomes variants. Discovery variants that were not in line with this reference were discarded. The processed LOO summary statistics were taken as input for the LDpred 0.9 softwareV1. For estimating the target LD structure, we took a selection of 2,500 unrelated individuals in the NTR database and selected a set of well-imputed variants in the NTR sample (21,53). The parameter ld_radius was set by dividing the number of variants in common (from the output of the coordination step) by 12000. For the coordination step we provided the median sample size as input value for N. For the LDpred step we applied the following thresholds for fraction of variants with a non-zero effect (in addition to the default infinitesimal model): --PS=0.5, 0.3, 0.2, 0.1, 0.05, 0.01, 0.005, 0.003, and 0.001.

In addition, we calculated a PGS in NTR based on 26 significant SNPs, that were also present in the Human Genome Diversity Project and the 1000 Genomes Project, from the most recent DZT GWAS (Table 2 from Mbarek et al., 2024). SNPs were selected from the LOO summary statistics and the PGS were calculated using the --score function of PLINK version 1.90.

#### MoBa

The complete genotyping and imputation information from MoBa can be found in Data S1. In MoBa we calculated the PGS in a similar way as in NTR. The full GWAS summary statistics, including the NTR, were taken as input for the LDpred 0.9 software. For estimating the target LD structure, the same sample was used (21,53). The parameter *ld_radius* was set by dividing the number of variants in common (from the output of the coordination step) by 12,000. Again, for the coordination step we provided the median sample size as input value for N. For the LDpred step we applied the 0.001 threshold for fraction of variants with non-zero effect. We chose to only include the 0.001 threshold for MoBa, based on the variance explained results from the NTR PGSs.

### Statistical methods

#### Polygenicity estimation

We employed the GENESIS tool as described by Zhang et al., 2018, for estimating the genetic effect-size distribution based on the summary statistics of the 2024 GWAMA of DZT (6,23). We first tested whether a two-components model (M2) or a three-components model (M3) as described in Zhang et al. (2018). We also estimated the proportion of SNPs with nonzero effects as an indication of the polygenicity in the SBayesS tool (24).

#### Variance explained by PGS on the liability scale

All analyses were executed in R4.3.2. For each set of weighted effect sizes (infinitesimal model and additional threshold, see above) we calculated the NTR PGSs over all genotype data sets. To select the optimal threshold value of the fraction of variants with non-zero effect we calculated the variance in DZT explained on the liability scale in 1,635 mothers of naturally conceived DZ twins and 2,797 controls employing the methods described by Lee et al. (54).

#### Odds ratios

For all 7,178 participants of the NTR, we determined in which decile their PGS was located. We then performed a logistic regression (*glm* function) for being a mother of a naturally conceived DZT and separately for being a MAR DZT mother on the PGS deciles. From this regression we determined the odds ratios and corresponding 95% confidence interval for each of the deciles using the *exp* function in rbase.

#### PGSs comparisons in NTR and MoBa

We performed logistic regression with PGS as predictor and case-control as outcome variable, corrected for relatedness by adding the family structure as a cluster (function *geeglm)* as both the NTR and MoBa sample included sister pairs. To correct for population stratification and other technical biases, the first 10 PCs and the genotyping platforms (as dummy variables) were added to the models as covariates. To investigate the effect of DZT related traits, we performed a second round of analyses that included the covariates maternal age at birth, height, BMI, smoking, and reproductive history (15).

We performed the following series of analyses in NTR and MoBa. Bonferroni adjusted significance thresholds were 7.1 × 10^-3^ (0.05 / 7) in NTR and 0.01 (0.05 /5) in MoBa.

1. Mothers of naturally conceived DZ twins compared to controls. In NTR controls are fathers of twins (non-twins themselves) and in MoBa controls were mothers of naturally conceived singletons.
2. Mothers of naturally conceived DZ twins compared to mothers of naturally conceived MZ twins.
3. Mothers of naturally conceived DZ twins compared to mothers of MAR DZ twins. We a further stratified the MAR DZ mothers in mothers who received OI treatment and other MAR mothers in the NTR.
4. Controls compared to mothers of naturally conceived MZ twins and mothers of MAR MZ twins.
5. In MoBa only: Mothers of naturally conceived singletons compared to mothers of MAR singletons.
6. Regression analyses with the PGS in mothers of naturally conceived twins (NTR) and singletons mothers (MoBa), and the TTP. TTP was analyzed both as a continuous variable and as a categorical variable stratified per months.

#### Genetic correlations

We applied LD Score Regression (LDSC; version v1.0.1; (22,55)) to calculate the genetic correlation of DZT with number of children ever born (NEB) and childlessness (CL) (56), PCOS (41), anovulatory infertility (28), endometriosis (40), miscarriages (57), and male infertility (58). The summary statistics of endometriosis were downloaded from the GWAS catalog while the other summary statistics were made available by the authors of these studies. Before the genetic correlations were calculated, the SNPs for each of the summary statistics were merged with the w_hm3.snplist, to ensure that the alleles listed in the summary statistics files match, which was downloaded from the LDSC wiki (https://github.com/bulik/ldsc/wiki/).

## Data Availability

The data of the NTR may be requested through the NTR data access committee (https://tweelingenregister.vu.nl/information_for_researchers/working-with ntr-data).
Data from the MoBa and the MBRN used in this study are managed by the national health register holders in Norway (Norwegian Institute of public health) and can be made available to researchers, provided approval from the Regional Committees for Medical and Health Research Ethics (REC), compliance with the EU General Data Protection Regulation (GDPR) and approval from the data owners. The consent given by the participants does allow storage of data on an individual level in repositories or journals. Researchers who want access to data sets for replication should apply through www.helsedata.no. Access to data sets requires approval from The Regional Committee for Medical and Health Research Ethics in Norway and an agreement with MoBa. All code can be made available from the corresponding author on request.

## Acknowledgements

We warmly thank all participant of the NTR and MoBa and all previous employees who contributed to the data collection and preparation.

NH is supported by the Royal Netherlands Academy of Science Professor Award (PAH/6635) to DIB. JvD is supported by Netherlands Organization for Scientific Research (NWO) Large Scale infrastructures, X-omics (184.034.019). The NTR is supported by multiple grants from the NWO and Medical Research (ZonMW): NTR Repository (NWO 480-15-001/674); CID Gravitation Program of the Dutch Ministry of Education, Culture, and Science and the Netherlands Organization for Scientific Research (NWO 0240-001-003). Genetic influences on stability and change in psychopathology from childhood to young adulthood (ZonMw 912- 10-020); Twin family database for behavior genomics studies (NWO 480-04-004); Twin research focusing on behavior (NWO 400-05-717); the European Science Council (ERC) Genetics of Mental Illness (ERC Advanced, 230374); Developmental trajectories of psychopathology (NIMH 1RC2 MH089995).

This work was partly funded by the Research Council of Norway (320656) and through its Centres of Excellence funding scheme (262700; CMP, SEH and JRH). We thank the Norwegian Institute of Public Health (NIPH) for generating high-quality genomic data. This research is part of the HARVEST collaboration, supported by the Research Council of Norway (229624). We also thank the NORMENT Centre for providing genotype data, funded by the Research Council of Norway (223273), South East Norway Versjon 7.0 3 Health Authorities and Stiftelsen Kristian Gerhard Jebsen. We further thank the Center for Diabetes Research, the University of Bergen for providing genotype data and performing quality control and imputation of the data funded by the ERC, Stiftelsen Kristian Gerhard Jebsen, Trond Mohn Foundation, the Research Council of Norway, the Novo Nordisk Foundation, the University of Bergen, and the Western Norway Health Authorities. The Norwegian Mother, Father, and Child Cohort Study is supported by the Norwegian Ministry of Health and Care Services and the Ministry of Education and Research. We are grateful to all the participating families who take part in this ongoing cohort study.

## Author contributions

NH, HM, GW, DIB, JRH, NL, NGM and CBL have contributed to the conceptualization of the study. NH, LL, GW, JJH, RP and CMP prepared the data for inclusion. NH and CMP have performed the analyses and NH was responsible for the first draft of the manuscript. RP, JvD, JJH, JRH, GW and DIB provided supervision. CMP, RP, HM, NL,VM, LL, JvD, ECC, JJB, EAE, NGM, GW, SH, JRH, JJH and DIB provided feedback and revision on the manuscript. All authors support the submitted version of the manuscript.

## Web resources

GENESIS: https://github.com/yandorazhang/GENESIS SBayesS: https://cnsgenomics.com/software/gctb/

GWAS Catalog: DZT: 27132594. Endometriosis: 36914876.

## Data and code availability

The data of the NTR may be requested through the NTR data access committee (https://tweelingenregister.vu.nl/information_for_researchers/working-with ntr-data).

Data from the MoBa and the MBRN used in this study are managed by the national health register holders in Norway (Norwegian Institute of public health) and can be made available to researchers, provided approval from the Regional Committees for Medical and Health Research Ethics (REC), compliance with the EU General Data Protection Regulation (GDPR) and approval from the data owners. The consent given by the participants does allow storage of data on an individual level in repositories or journals. Researchers who want access to data sets for replication should apply through www.helsedata.no. Access to data sets requires approval from The Regional Committee for Medical and Health Research Ethics in Norway and an agreement with MoBa. All code can be mase available from the corresponding author on request.

