## Supplementary data for "Polygenic Scores for Dizygotic Twinning: Insights into the Genetic Architecture of Female Fertility"

**Supplemental data**

**Figure S1**: QQplots of the A) two-components (M2) and the B) three-component (M3) models of the GENESIS tool applied to the dizygotic twinning (DZT) GWAS summary statistics.

**Figure S2:**  The predicted % of the total estimated SNP heritability of dizygotic twinning (DZT) explained with varying sample size of the GWAS. The sample size include a ratio of cases and controls of 1:1.

**Figure S3:** The mean PGS for dizygotic (DZ) twinning in controls, mothers of naturally conceived monozygotic (MZ) twins and mothers of MZ twins that received medically assisted reproduction (MAR) treatments from **A)** the Netherlands Twin Register (NTR) and **B)** the Norwegian Mother, Children, and Father Cohort Study (MoBa).

**Figure S4:** The mean PGS for dizygotic (DZ) twinning in mothers of naturally conceived singletons (controls) and mothers of singletons who received medically assisted reproduction (MAR) treatments from the Norwegian Mother, Children, and Father Cohort Study (MoBa).

**Figure S5:** The mean PGS for dizygotic (DZ) twinning in mothers of twins that are identified as ethnic outliers in the Netherlands twin register (NTR). MAR = Medically assisted reproduction

**Table S1:** The phenotypic variance explained by each PGS calculation based on the different fractions of SNPs included in the PGS.

**Table S2:** The comparison of the polygenic scores (PGS) for dizygotic (DZ) twinning in mothers of DZ with controls and other mothers of twins from the Netherlands Twin Register (NTR), including the correction for the covariates maternal age, age at firth birth, height, BMI before pregnancy, number of older children and smoking before the twin pregnancy.. The controls consist of fathers of twins. All the analyses are shown with the mothers of naturally conceived DZ twins as the reference group. MAR = Medically assisted reproduction; OI = Ovulation induction.

**Table S3:** The comparison of the polygenic score for dizygotic (DZ) twinning in mothers of DZ twins with mothers of MZ twins and singletons from the Norwegian Mother, Children, and Father (MoBa). The controls include the mothers of naturally conceived singletons. These analyses include the correction for the covariates maternal age, age at firth birth, height, BMI before pregnancy, number of older children and smoking before the twin pregnancy. All the results are shown with the mothers of naturally conceived DZ twins as the reference group. MAR = Medically assisted reproduction.

**Table S4:** Results of the comparison of the polygenic score for dizygotic (DZ) twinning in controls or monozygotic (MZ) twins from the Netherlands twin register. The controls consist of fathers of twins. All the analyses are showed with the controls as reference group. MAR = Medically assisted reproduction

**Table S5:** Results of the comparison of the polygenic score for dizygotic (DZ) twinning in mothers of singletons from the Norwegian Mother, Children, and Father (MoBa). The controls contain the mothers of naturally conceived singletons. The controls consist of fathers of twins. All the analyses are showed with the controls as reference group. MAR = Medically assisted reproduction

**Table S6:** Results of the regression and factor analyses of the time to pregnancy (TTP) in month and the PGS for DZT in mothers of naturally conceived twins from the NTR. The analyses were performed both with TTP as a continuous variable (TTP all) and as factor per category, with 0-2 months till pregnancy as reference group.

**Table S7:** Results of the regression and factor analyses of the time to pregnancy (TTP) in month and the PGS for DZT in mothers of naturally conceived singletons of the from the Norwegian Mother, Children, and Father (MoBa) cohort. The analyses were performed both with TTP as a continuous variable (TTP all) and as factor per month with mothers of an unplanned pregnancy as reference group.

**Data S1:** The Genotyping and imputation information of A) the Netherlands twin register (NTR) and B) the Norwegian Mother, Children, and Father (MoBa) cohort.

**Figure S1**: QQplots of the A) two-components (M2) and the B) three-component (M3) models of the GENESIS tool applied to the dizygotic twinning (DZT) GWAS summary statistics.

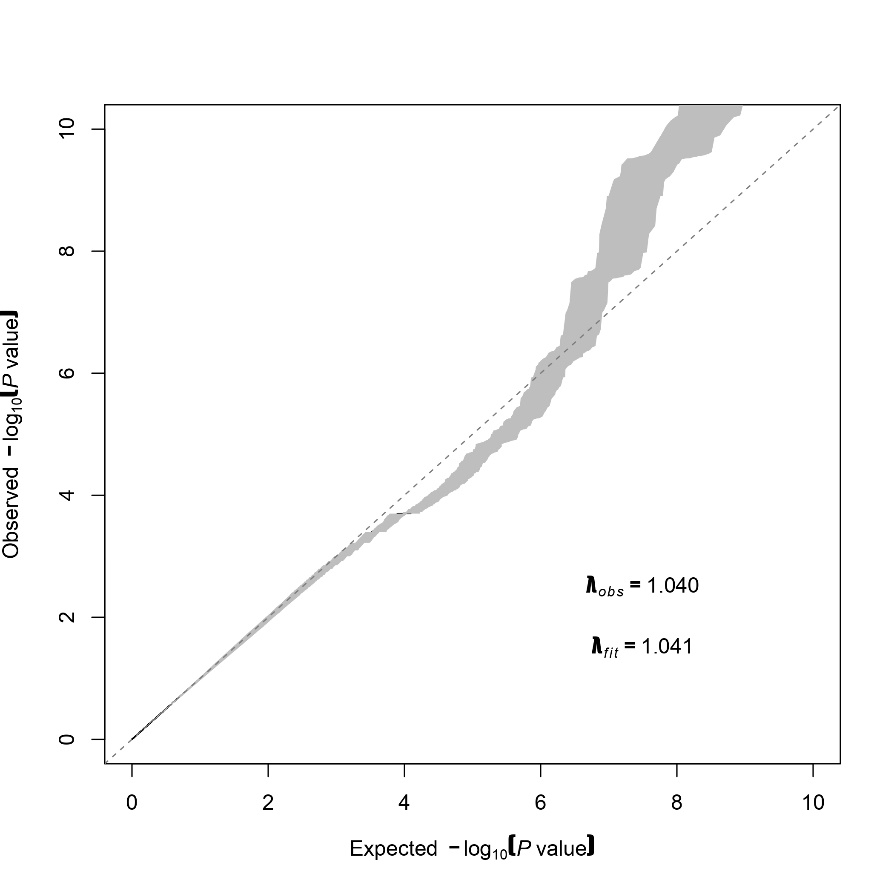

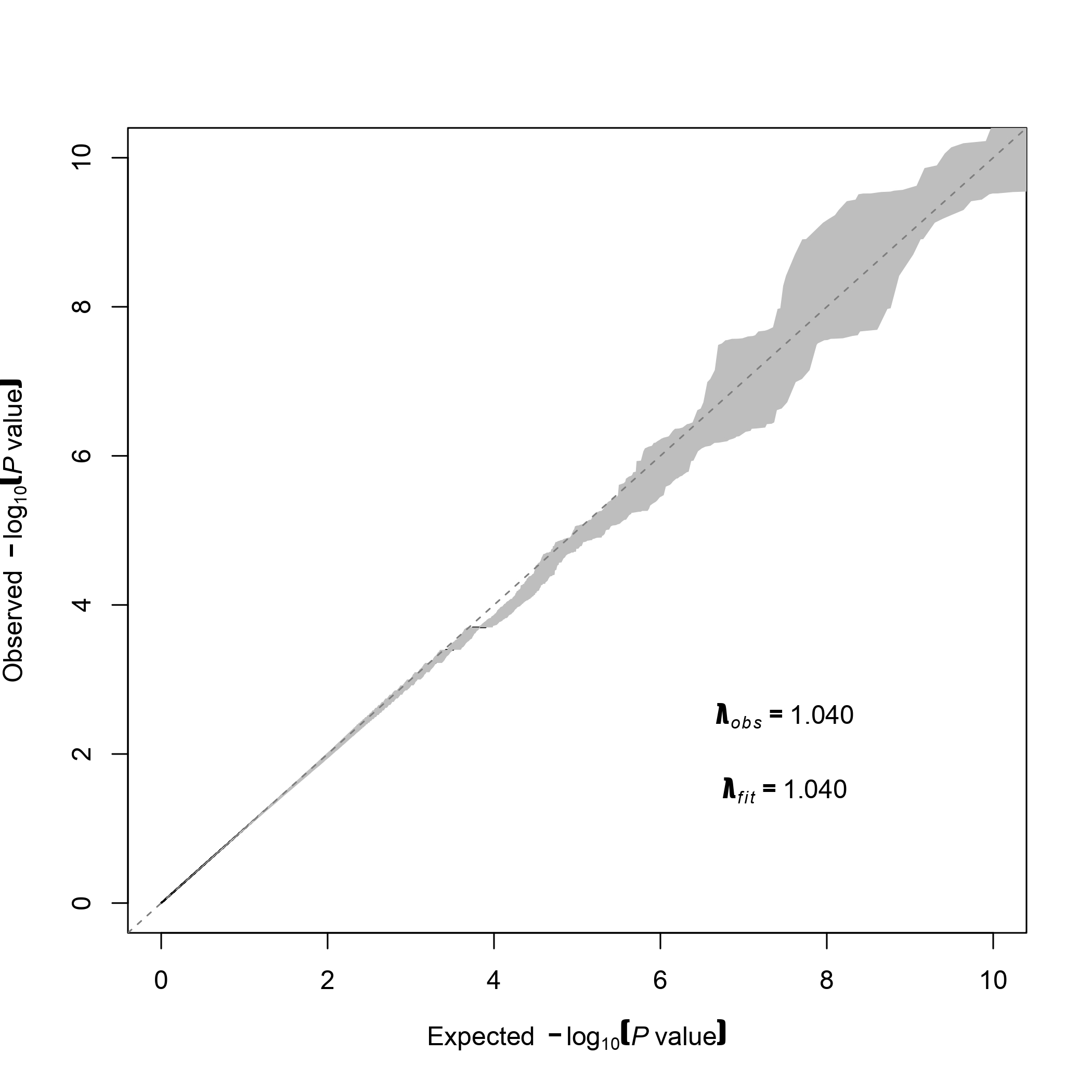

**B**

**A**

**Figure S2:**  The predicted % of the total estimated SNP heritability of dizygotic twinning (DZT) explained with varying sample size of the GWAS. The sample size include a ratio of cases and controls of 1:1.

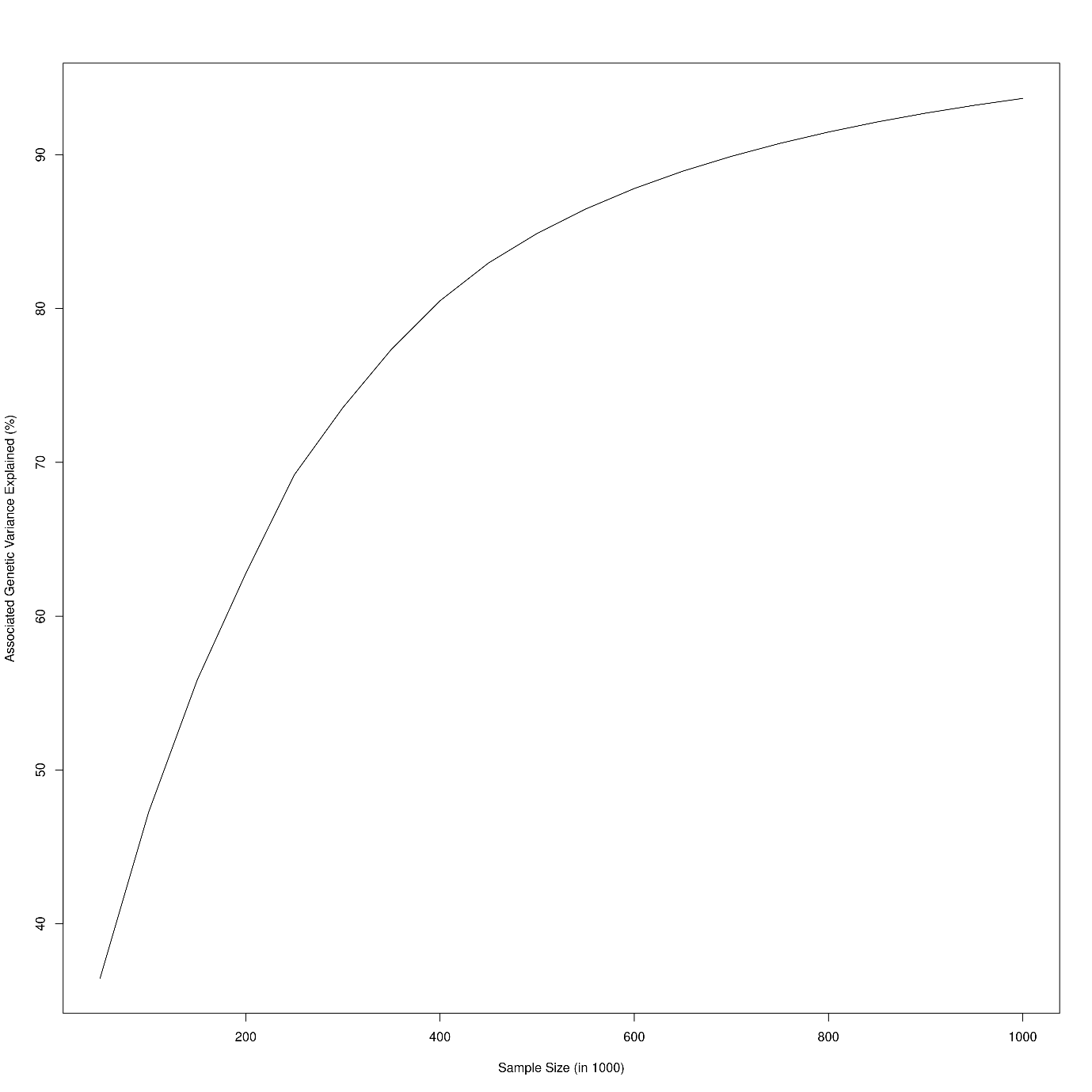

**Figure S3:** The mean PGS for dizygotic (DZ) twinning in controls, mothers of naturally conceived monozygotic (MZ) twins and mothers of MZ twins that received medically assisted reproduction (MAR) treatments from **A)** the Netherlands Twin Register (NTR) and **B)** the Norwegian Mother, Children, and Father Cohort Study (MoBa).

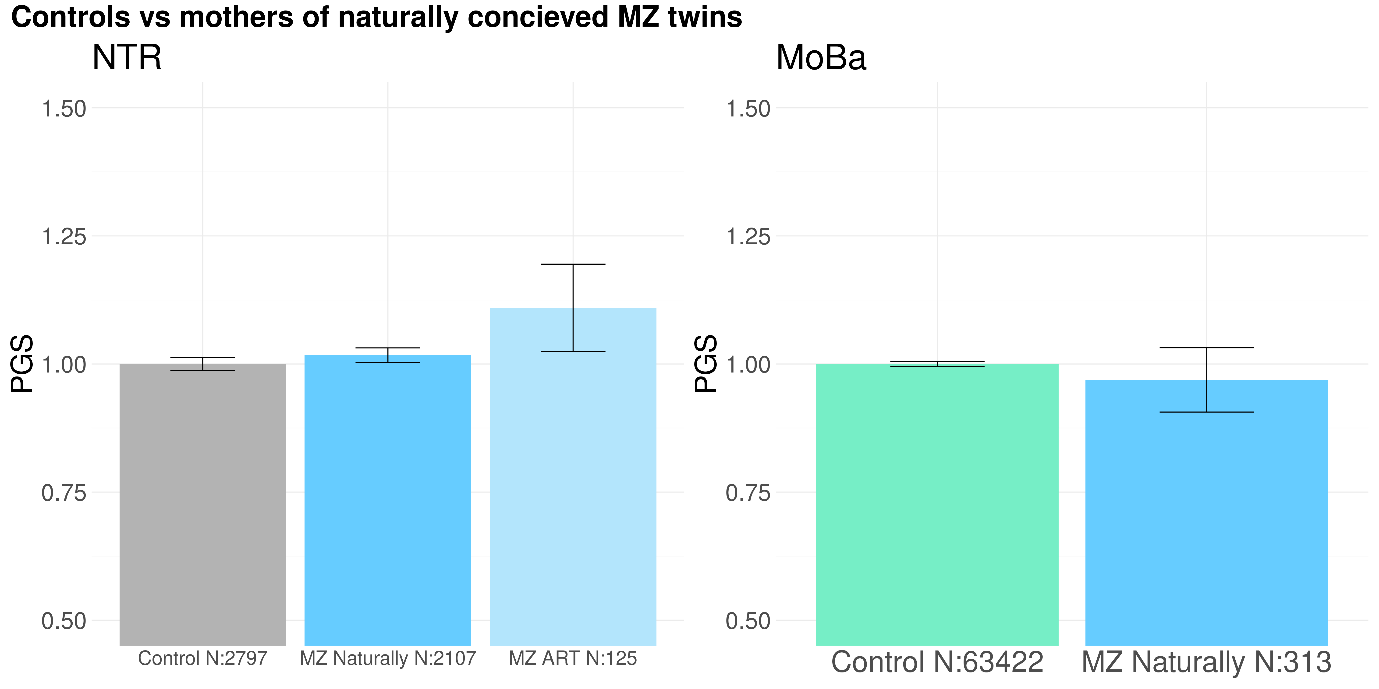

**Figure S4:** The mean PGS for dizygotic (DZ) twinning in mothers of naturally conceived singletons (controls) and mothers of singletons who received medically assisted reproduction (MAR) treatments from the Norwegian Mother, Children, and Father Cohort Study (MoBa).

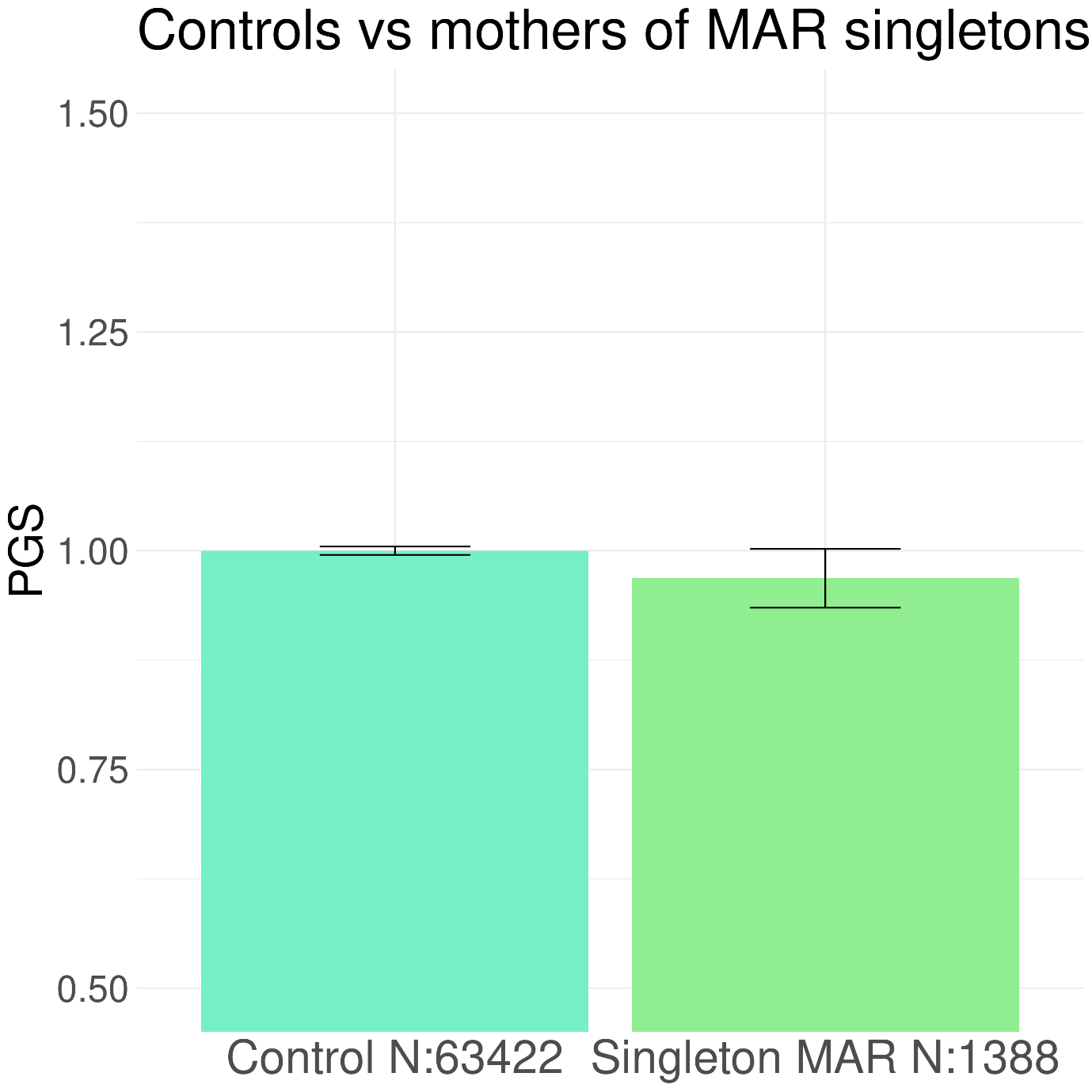

**Figure S5:** The mean PGS for dizygotic (DZ) twinning in mothers of twins that are identified as ethnic outliers in the Netherlands twin register (NTR). The bars and standard error lines are standardized on the mean PGS from the mothers of naturally conceived DZ twins. MAR = Medically assisted reproduction

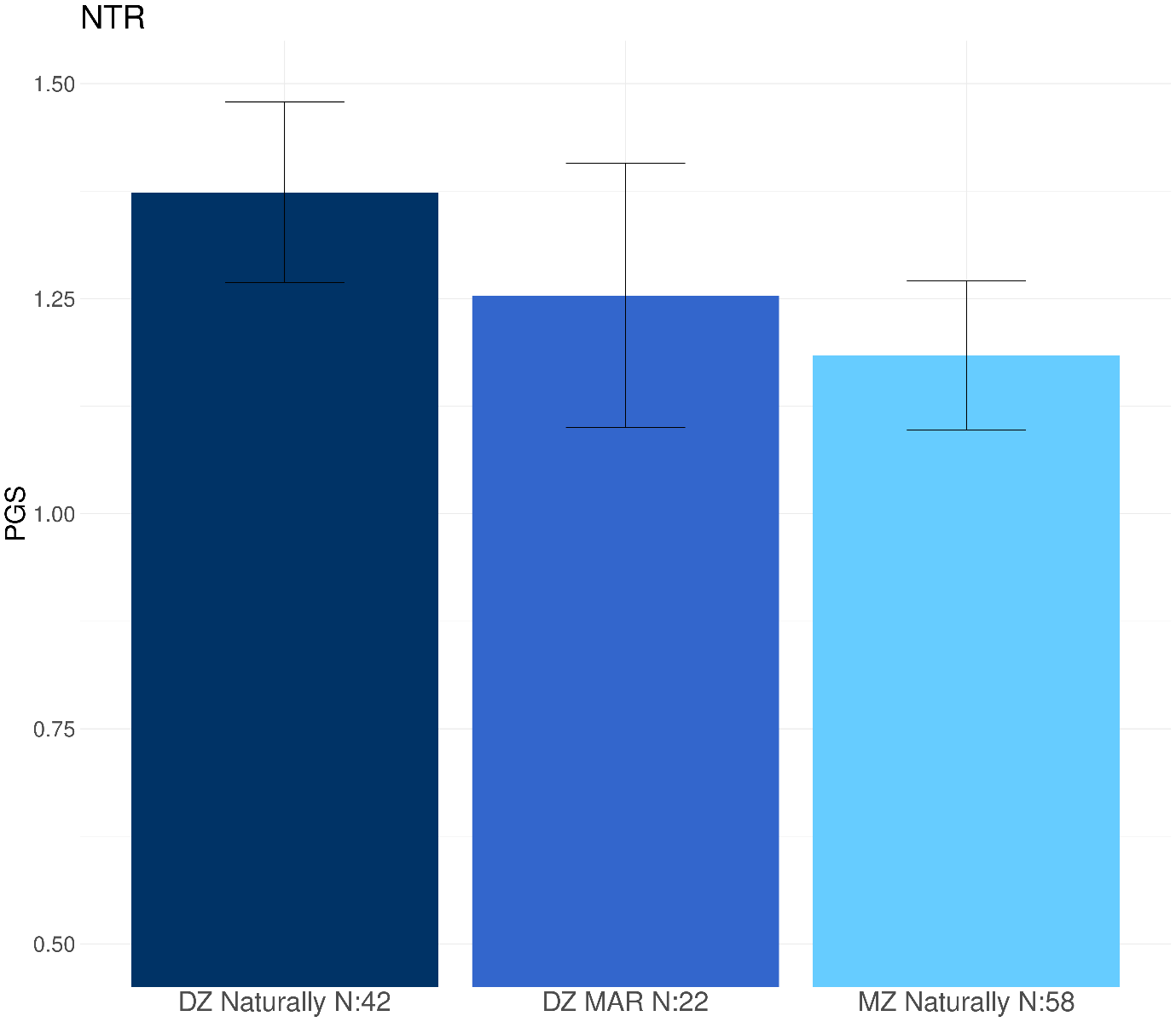

**Table S1:** the phenotypic variance explained by each PGS calculation based on the different fractions of SNPs included in the PGS.

| Fraction of SNPs | Variance explained % |
| --- | --- |
| All | 0.79% |
| 0.5 | 0.86% |
| 0.3 | 0.86% |
| 0.2 | 0.87% |
| 0.1 | 0.87% |
| 0.05 | 0.88% |
| 0.03 | 0.90% |
| 0.01 | 1.00% |
| 0.005 | 1.10% |
| 0.003 | 1.22% |
| 0.001 | 1.60% |
| Top 26 SNPS | 0.71% |

**Table S2:** The comparison of the polygenic scores (PGS) for dizygotic (DZ) twinning in mothers of DZ with controls and other mothers of twins from the Netherlands Twin Register (NTR), including the correction for the covariates maternal age, age at firth birth, height, BMI before pregnancy, number of older children and smoking before the twin pregnancy.. The controls consist of fathers of twins. All the analyses are shown with the mothers of naturally conceived DZ twins as the reference group. MAR = Medically assisted reproduction; OI = Ovulation induction.

| Group | N | Mean PGS | P value | Beta (95% CI) |
| --- | --- | --- | --- | --- |
| DZ Naturally | 848 | 3.75x10^-9^ | - | - |
| DZ MAR | 303 | 3.15x10^-9^ | *3.22x10^-5^* | -1.47x10^8^ (-2.17x10^8^- -7.61x10^7^) |
| DZ OI | 147 | 2.81x10^-9^ | *1.20x10^-6^* | -2.29x10^8^ (-1.34x10^8^- -3.23x10^8^) |
| DZ rest MAR | 156 | 3.49x10^-9^ | 0.177 | -6.36x10^7^ (-1.58x10^8^ - 3.07x10^7^) |
| MZ Naturally | 1233 | 3.29x10^-9^ | *4.54x10^-7^* | -1.10x10^8^ (-6.62x10^7^- -1.53x10^8^) |
| MZ MAR | 85 | 3.59x10^-9^ | 0.0834 | -1.07x10^8^ (-2.31x10^8^- 1.66x10^7^) |

| Group | N | Mean PGS | P value | Beta (95% CI) |
| --- | --- | --- | --- | --- |
| Controls | 63422 | 1.90x10^-9^ | *1.30x10^-4^* | -6.19x10^7^ (-9.43x10^7^ - -2.95x10^7^) |
| DZ Naturally | 658 | 2.25x10^-9^ | *-* | - |
| DZ MAR | 328 | 1.74x10^-9^ | *0.0015* | -1.17x10^8^ (-1.90x10^8^- -4.31x10^7^) |
| MZ Naturally | 313 | 1.84x10^-9^ | 0.024 | -7.86x10^7^ (-1.53x10^8^- -1.48x10^6^) |
| Singletons MAR | 1388 | 1.84x10^-9^ | *2.10 x10^-4^* | -7.47x10^7^ (-1.15x10^8^- -3.43x10^7^) |

**Table S3:** The comparison of the polygenic score for dizygotic (DZ) twinning in mothers of DZ twins with mothers of MZ twins and singletons from the Norwegian Mother, Children, and Father (MoBa). The controls include the mothers of naturally conceived singletons. These analyses include the correction for the covariates maternal age, age at firth birth, height, BMI before pregnancy, number of older children and smoking before the twin pregnancy. All the results are shown with the mothers of naturally conceived DZ twins as the reference group. MAR = Medically assisted reproduction.

**Table S4:** Results of the comparison of the polygenic score for dizygotic (DZ) twinning in controls or monozygotic (MZ) twins from the Netherlands twin register. The controls consist of fathers of twins. All the analyses are showed with the controls as reference group. MAR = Medically assisted reproduction

| Group | N | Mean PGS | P value | Beta (95% CI) |
| --- | --- | --- | --- | --- |
| Controls | 2797 | 3.23x10^-9^ | - | - |
| MZ Naturally | 2107 | 3.29x10^-9^ | 0.350 | 1.27x10^7^ (-1.45x10^7^- 4.00x10^7^) |
| MZ MAR | 125 | 3.59x10^-9^ | 0.185 | 7.61x10^7^ (-3.86x10^7^- 1.91x10^8^) |

**Table S5:** Results of the comparison of the polygenic score for dizygotic (DZ) twinning in mothers of singletons from the Norwegian Mother, Children, and Father (MoBa). The controls contain the mothers of naturally conceived singletons. The controls consist mothers of naturally conceived singletons. All the analyses are showed with the controls as reference group. MAR = Medically assisted reproduction

| Group | N | Mean PGS | P value | Beta (95% CI) |
| --- | --- | --- | --- | --- |
| Controls | 63422 | 1.90x10^-9^ | - | - |
| MZ Naturally | 313 | 1.84x10^-9^ | 0.84 | 5.04x10^7^ (-4.42x10^8^ – 5.43x10^8^) |
| Singletons MAR | 1388 | 1.84x10^-9^ | 0.32 | -1.23x10^7^ (-3.67x10^7^- 1.21x10^7^) |

**Table S6:** Results of the regression and factor analyses of the time to pregnancy (TTP) in month and the PGS for DZT in mothers of naturally conceived twins from the NTR. The analyses were performed both with TTP as a continuous variable (TTP all) and as factor per category, with 0-2 months till pregnancy as reference group.

|  | N | P value | Beta (95% CI) |
| --- | --- | --- | --- |
| TTP all | 2298 | 0.280 | -1.18e-11 (-3.36e-11 - 1.00e-11) |
| TTP 3-5 months | 569 | 0.394 | 9.52e-11 (-1.28e-10 - 3.19e-10) |
| TTP 6-12 months | 336 | 0.258 | -1.48e-10 (-4.11e-10 - 1.14e-10) |
| TTP 12 or more months | 302 | 0.550 | -8.15e-11 (-3.54e-10 - 1.91e-10) |

**Table S7:** Results of the regression and factor analyses of the time to pregnancy (TTP) in month and the PGS for DZT in mothers of naturally conceived singletons of the from the Norwegian Mother, Children, and Father (MoBa) cohort. The analyses were performed both with TTP as a continuous variable (TTP all) and as factor per month with mothers of an unplanned pregnancy as reference group.

|  | N | P value | Beta (95% CI) |
| --- | --- | --- | --- |
| TTP all | 42285 | *0.0036* | -1.20e-12 (-3.76e-13- -2.03e-12) |
| TTP 1 month or less | 12180 | 0.986 | 7.54e-14 (-8.32e-12 - 8.47e-12) |
| TTP 2 months | 14326 | 0.189 | -5.33e-12 (-1.34e-11 - 2.78e-12) |
| TTP 3 months | 1087 | 0.557 | -6.06e-12 (-2.67e-11- 1.46e-11) |
| TTP 4 months | 2837 | 0.0938 | -1.12e-11 (-2.46e-11 - 2.16e-12) |
| TTP 5 months | 1993 | 0.537 | -4.91e-12 (-2.08e-11 - 1.10e-11) |
| TTP 6 months | 2250 | 0.669 | 3.23e-12 (-1.19e-11 - 1.83e-11) |
| TTP 7 months | 984 | 0.159 | -1.57e-11 (-3.80e-11 - 6.58e-12) |
| TTP 8 months | 843 | 0.784 | 3.21e-12 (-2.02e-11 - 2.66e-11) |
| TTP 9 months | 670 | 0.431 | -9.81e-12 (-3.47e-11 - 1.51e-11) |
| TTP 10 months | 666 | 0.194 | -1.73e-11 (-4.39e-11 - 9.34e-12) |
| TTP 11 months | 385 | 0.241 | -1.95e-11 (-5.29e-11 - 1.38e-11) |
| TTP 12 months or more | 4064 | *0.0138* | -1.48e-11 (-2.78e-12 - -2.68e-11) |

**Data S1**

***Genotyping and imputation***

*NTR*

We used version 18 of the NTR genotyping data. Genotyping was conducted on three SNP array platforms: Affymetrix 6.0 (N=10,377), Affymetrix Axiom (N=3536), and Illumina GSA NTR array (N=20,060). Genotype calling followed the manufacturer protocols and white papers. DNA samples were checked for gender mismatches, heterozygosity and Identity by Descent (IBD) mismatches in comparison to the known family structure. Each sample needed to have a call rate of at least 90% and at least 80% of the genotypes needed to be present on each chromosome. SNP quality control was based on the following filters applied in each platform: call rate over 95%, Hardy-Weinberg equilibrium (HWE) p-value over 0.0001, minor allele frequency over 0.01 and Mendelian as well as genotyping error rate less than 1%. This led to the final set of 3,644 individuals with 534,405 SNPs on Axiom, 9,049 individuals with 537,992 SNPs on Affymetrix 6.0 and 16,276 individuals with 481,898 SNPs on Illumina GSA, a total of 28,969 individuals. After strand and name alignment of the data, removing palindromic SNPS with a MAF>0.30, the genotype data were imputed with Beagle 5.4 against the 1000 Genomes, and a combined HRC 1.1 (Ega version) plus GoNL reference panel (Boomsma et al., 2013; McCarthy et al., 2016).

Twenty 1000 Genomes projected principal components (PC) for the genotype data were calculated with the EIGENSTRAT smartpca tool (Kraus et al., 2013). We selected the SNPs that passed quality control (QC) and were present in one of the three platforms from the 1000 Genomes imputed data (as the overlap between platforms is too small to take only genotyped SNPs). SNPs were then filtered to have MAF>0.05, HWE p > 0.001, call rate > 0.98, Mendelian error rate < 1% and imputation info>=90%. These SNPs were subsequently pruned for linkage disequilibrium (LD) with Plink version 1.9 (option –indep 50 5 2) and SNPs in long range LD blocks were removed as described in (Purcell et al., 2007)Abdellaoui et al., 2013). This left 110,558 SNPs for analysis. From the 1000 Genomes reference panel all samples with the same SNPs were selected and then merged with the NTR data. Subsequently PCs were calculated in the 1000 Genomes set, and then projected upon the NTR data with the smartpca software.

*MoBa*

Complete details of MoBa genotyping, pre-imputation QC, phasing, imputation, and post-imputation QC are described elsewhere (Corfield et al., 2024). Here we will provide a summary. In total, 238,001 MoBa samples were genotyped in 24 genotyping batches with varying selection criteria, genotyping arrays and genotyping centers. SNPs were temporarily removed based on the following criteria: call rate < 95%; out of HWE at p < 1.00 ⨉ 10-3; and MAF < 1%. Individuals were temporarily removed with call rate ≤ 95%. Principal component analysis (PCA) was performed with 1000 Genomes phase 1 (N = 1,083 unrelated individuals) to identify the subpopulations (Corfield et al., 2022). To perform the PCA, LD pruning was performed and SNPs in long-range high LD regions were removed before merging the batch genotypes with those of the 1000 Genomes data. PCs were first estimated within founders (based on reported information). The non-founders, all individuals with at least one reported parent, were then projected into the PC space of founders. Individuals were then assigned to European, Asian, and African core subpopulations based on visual inspection using the first seven PCs.

Within the subpopulation another QC round was performed checking SNPs on: SNPs with MAF < 0.5%; SNPs with call rate < 95%; SNPs with, call rate < 98%; SNPs and individuals with a call rate < 98%; and SNPs out of HWE at p < 1.00 ⨉ 10-6. Individuals were checked on sex mismatches, a relatedness check with IBD by employing KING version 2.2.5 and heterozygosity filtering was then performed with individual outliers removed if they were ± 3 standard deviations from the mean heterozygosity across all individuals using only autosomes. Additional ancestry outliers were removed based on PCA with the 1000 Genomes phase 1 unrelated data.

SNPs passing all rounds of QC were merged by genotyping array. The publicly available European Genome-Phenome Archive (Study ID EGAS00001001710) HRC release 1.1 data was used as the reference panel for both phasing and imputation. Post-imputation QC was performed in a single round following the QC by genotype array protocol for each imputation batch. The following thresholds were used for SNP removal: imputation quality score ≤ 0.8; MAF < 1%; call rate < 95%; HWE p-value < 1.00⨉ 10^-6^; discordant in true duplicates; > 1% ME; and association with genotype batch at a p-value < 5 ⨉ 10^-8^. The following checks were used for individual removal: call rate < 98%; ± 3 standard deviations from the mean heterozygosity across all individuals; the individual from each true duplicate pair with the lowest call rate; relatedness checks; cryptic relatedness; ME > 5% in families; ancestry outliers; and subpopulation outliers. Of the 235,412 successfully genotyped samples, 234,505 came from individuals still included in the study as of January 2019.
